# tame: An R package for identifying clusters of medication use based on dose, timing and type of medication

**DOI:** 10.1101/2024.06.24.24309427

**Authors:** Anna Laksafoss, Jan Wohlfahrt, Anders Hviid

**Author notes:** Corresponding Author: Anna Laksafoss.

## Abstract

Simplified exposure classifications, such as ever exposed versus never exposed, are commonly used in pharmacoepidemiology. However, this simplification may obscure complex use patterns relevant to researchers. We introduce tame, an R package that offers a novel method for classifying medication use patterns, capturing complexities such as timing, dose, and concurrent medication use in real-world data. The core innovation of tame is its bespoke distance measure, which identifies complex clusters in medication use and is highly adaptable, allowing customization based on the Anatomical Therapeutic Chemical (ATC) Classification System, medication timing, and dose. By prioritizing a robust distance measure, tame ensures accurate and meaningful clustering, enabling researchers to uncover intricate patterns within their data. The package also includes tools for visualizing and applying these clusters to new datasets. In a national Danish cohort study, tame identified nuanced antidepressant use patterns before and during pregnancy, demonstrating its capability to detect complex trends. tame is available on the Comprehensive R Archive Network at [https://CRAN.R-project.org/package=tame] under an MIT license, with a development version on GitHub at [https://github.com/Laksafoss/tame]. tame enhances medication use classification by detecting complex interactions and offering insights into real-world medication usage, thus improving stratification in epidemiological studies.

## INTRODUCTION

Developments in the field of machine learning, improvements in computing power, and the increasing richness of data now make it feasible to conduct more nuanced analyses of medication patterns. Central to these analyses is the construction of effective distance measures, which are critical in determining the success of clustering methods. A number of recent studies have explored data-driven approaches to exposure classification. For example, methods such as longitudinal k- means and latent class analysis have been applied to longitudinal medication data to classify exposure trajectories [1, 2, 3, 4], and these methods have been implemented in software such as the R-packages kml [5, 6] and lcmm [7]. However, these existing methods exhibit significant limitations.

Primarily, they focus exclusively on drug exposure timing, which restricts their ability to account for the full complexity of medication use patterns. By considering only the timing of drug exposure, these methods overlook other critical factors such as dose intensity and the chemical and therapeutic characteristics of the medications involved. This narrow focus limits their applicability, particularly in scenarios where the interplay between different medications and their specific attributes is crucial, such as in studies of drug safety and efficacy. Few developments have been made to approaches which learn from both exposure timing and the chemical and therapeutic characteristics of the medication(s) used [8]. To the authors’ knowledge, no publicly available software that learns from this combined information to identify real-world prescription drug use patterns exists.

In this paper, we present a novel distance measure explicitly designed for the clustering of medication use patterns, integrating exposure timing, dose intensity, and medication type based on Anatomical Therapeutic Clustering (ATC) codes. This distance measure is the foundation of our Hierarchical Cluster Analysis (HCA) implemented in the ‘medic()’ function in our package tame. HCA is a general and highly adaptable approach to cluster analysis that allows for the clustering of any data, provided that the user can specify the distance between any pair of observations. We developed this distance measure specifically for the clustering of based on three of the central characteristics of personal medication use: patterns of medication use by Anatomical Therapeutic Clustering (ATC) code, exposure timing, and exposure dose. The distance measure allows for flexible specification of the component parts – ATC code, timing and dose – and their relative importance. The method avoids unwanted data reduction or oversimplification of information by defining the distance measure on the person- and medication-specific level. Here, we apply the method to a Danish nationwide cohort of pregnancies with at least one antidepressant used 0-24 weeks prior to pregnancy onset in order to map Danish mothers’ use of antidepressants leading up to- and during pregnancy.

## IMPLEMENTATION

tame is implemented in R (R Development Core Team 2022) as a package and requires the following R packages: dplyr (≥1.1.0), fuzzyjoin, magrittr, purrr, Rfast, rlang, stats, stringr, tibble, tidyr, tidyselect, Rcpp (≥ 1.0.8). A development version is also available on GitHub [https://github.com/Laksafoss/tame] and collaborators are welcome to fork or make pull requests.

The tame package provides methods for clustering medication data by ATC codes, dose and timing, analysing and illustrating these clusters, and employing these learned clusters to new data in a similar format. These central functionalities are implemented in the functions ‘medic()’, ‘summary()’, and ‘employ()’, along with a few supporting functions. The package also includes a simulated dataset for demonstrating the code.

### Clustering Analysis

Our medication clustering method, implemented in the ‘medic()’ function, utilizes a novel distance measure within an agglomerative hierarchical cluster analysis (HCA) framework. The bespoke distance measure drives the clustering process by accurately quantifying the similarities between any two individual-level medication use profiles. The quality and effectiveness of the clustering hinge on this measure, making it the critical element of our methodology.

This distance measure is a composite that integrates multiple dimensions of medication information: dose intensity, timing of medication and the ATC classification system. Figure 1 provides an overview of this method, illustrating how the distance measure is constructed and applied within the clustering process to group individuals based on their medication use patterns.

**Figure 1.**
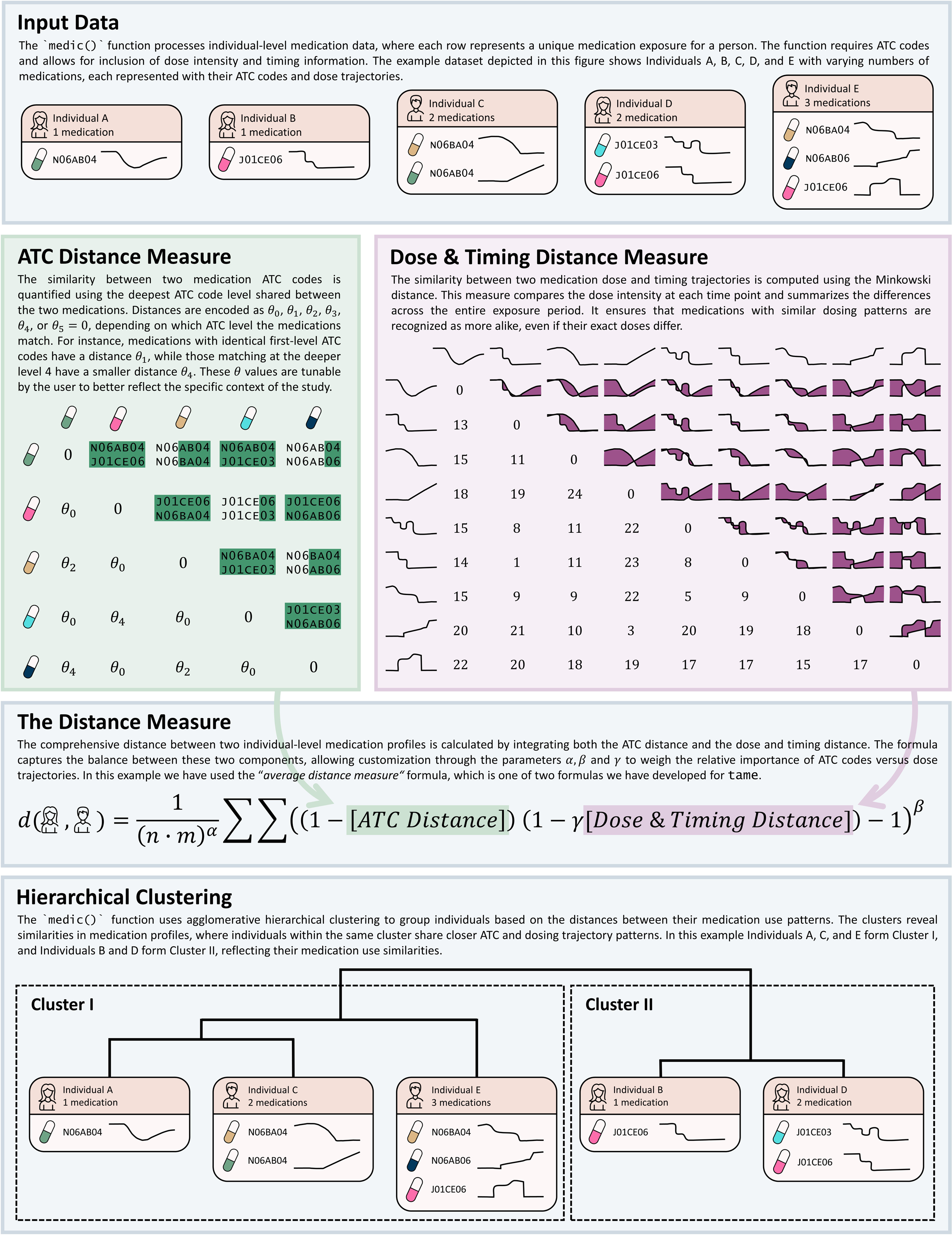
Overview of the ‘medic()’ function for quantifying and clustering individual-level medication profiles. The method calculates the similarity between medications using ATC codes and dosing trajectories, combining these into a comprehensive distance measure that accounts for both dose intensity and timing. Hierarchical clustering is then applied to group individuals based on these medication use patterns, revealing similarities in their profiles.

Difference in two individual-level medication profiles by dose intensity and timing of medication are measured by a dose trajectory distance measure. This distance measure compares the medication dose at each timepoint in the study and summarizes across time using the Minkowski distance. The mathematics behind the method are available in S1, and a mathematical glossary is available in S2 The ATC distance measure compares two medications by considering the levels of the medications ATC code. The distance between two medications is found by identifying the deepest ATC code level that is shared between the two medications. For example, sertraline N06AB06 and clomipramine N06AA04 are both antidepressants (N06A) and are identical at ATC level 3, but sertraline N06AB06 and oxycodone N02AA05 are only identical at ATC level 1, where they share the nervous system main group (N). Thus, the distance between sertraline N06AB06 and clomipramine N06AA04 is smaller than the distance between sertraline N06AB06 and oxycodone N02AA05. Note that if two medications have the same ATC codes, their distance is 0, and they are considered to be the same medication under this distance measure.

A wide range of parameters allow for the tuning of the dose trajectory distance measure, the ATC distance measure, and their relative importance. This flexibility ensures that the distance measure can be adapted to the specific context and research questions, preserving the complexity and richness of the underlying data. Detailed explanations and discussions of these tuning parameters and more can be found in S3.

While the method – like all clustering methods – is computationally intensive, these demands are justified by the need for accurate and meaningful clustering based on the detailed distance measure. For a technical discussion on these aspects of the implementation, see S4.

‘medic()’:Identifying clusters

The ‘medic()’ function is the central power house of the tame package. This function computes the distances between all pairs of individuals, runs the hierarchical clustering algorithm, and returns the clusters. See Figure 1 for a visual guide to the steps performed by the ‘medic()’ function, including how it calculates the composite distance measure and applies hierarchical clustering to group individuals based on their medication use profiles.

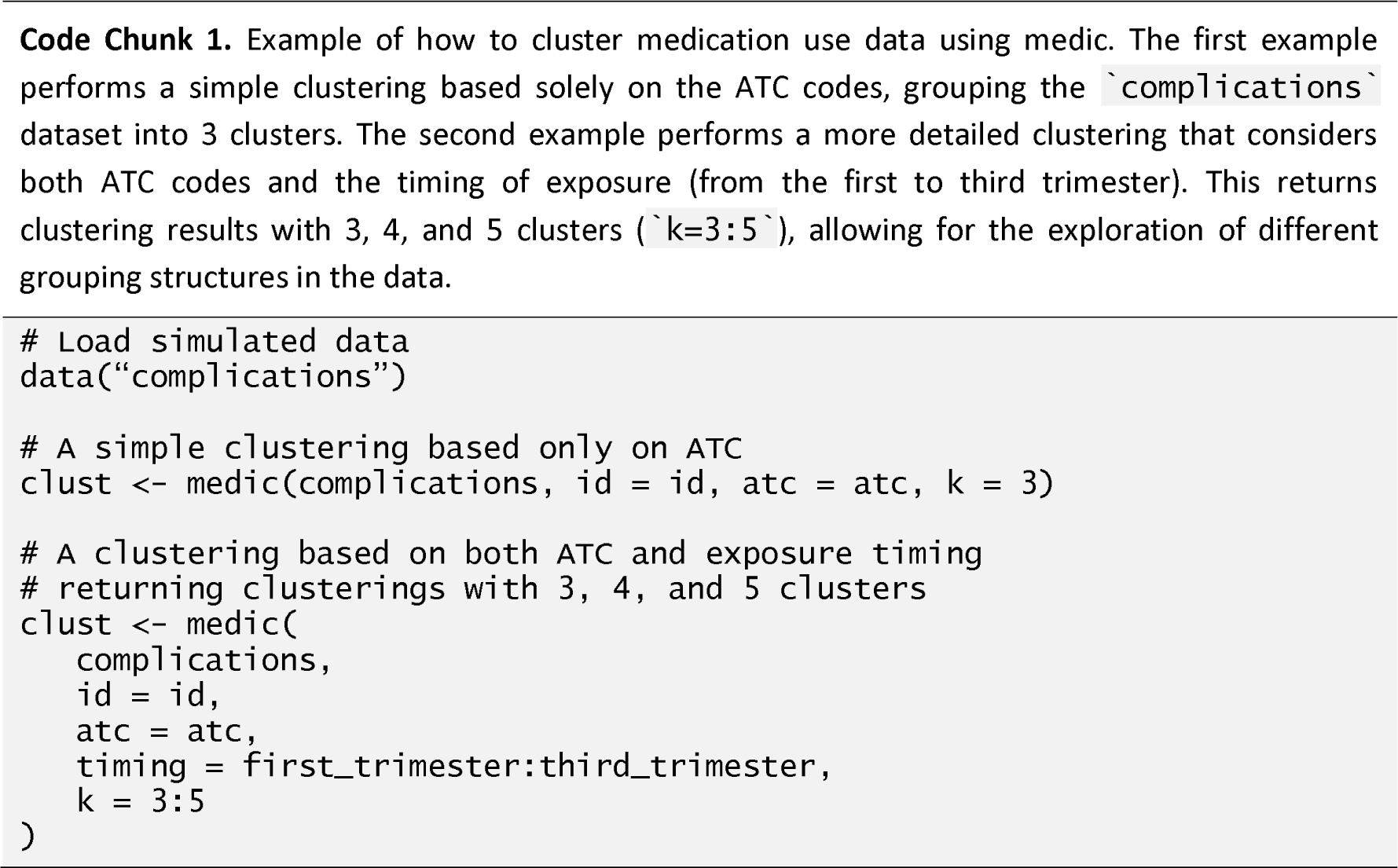

The function requires that the user provides a dataset where each row encodes personal medication patterns of each medication. Thus, a person exposed to 3 medications has 3 rows in the data. In S5 examples of how data might be structured can be found. Columns in this dataset naming the person identification variable, the medication ATC code variable, and, if available, one or more numerical variables encoding dose and/or exposure trajectory. The user specifies the desired number(s) of clusters with the function input ‘k’. Moreover, a number of function parameters can be used to tune the distance. A full discussion of these tuning parameters can be found in S3.

‘summary()’: Summarizing clusters

A wide range of cluster summarisation tools are implemented. Summary functions investigates different aspects of the clustering characteristics and have a corresponding plotting method. The list of summarisation approaches is given in the code documentation [9], and the latest version at time of publication can be found in S7. As an example, Figure 2 has been made using these summary tools.

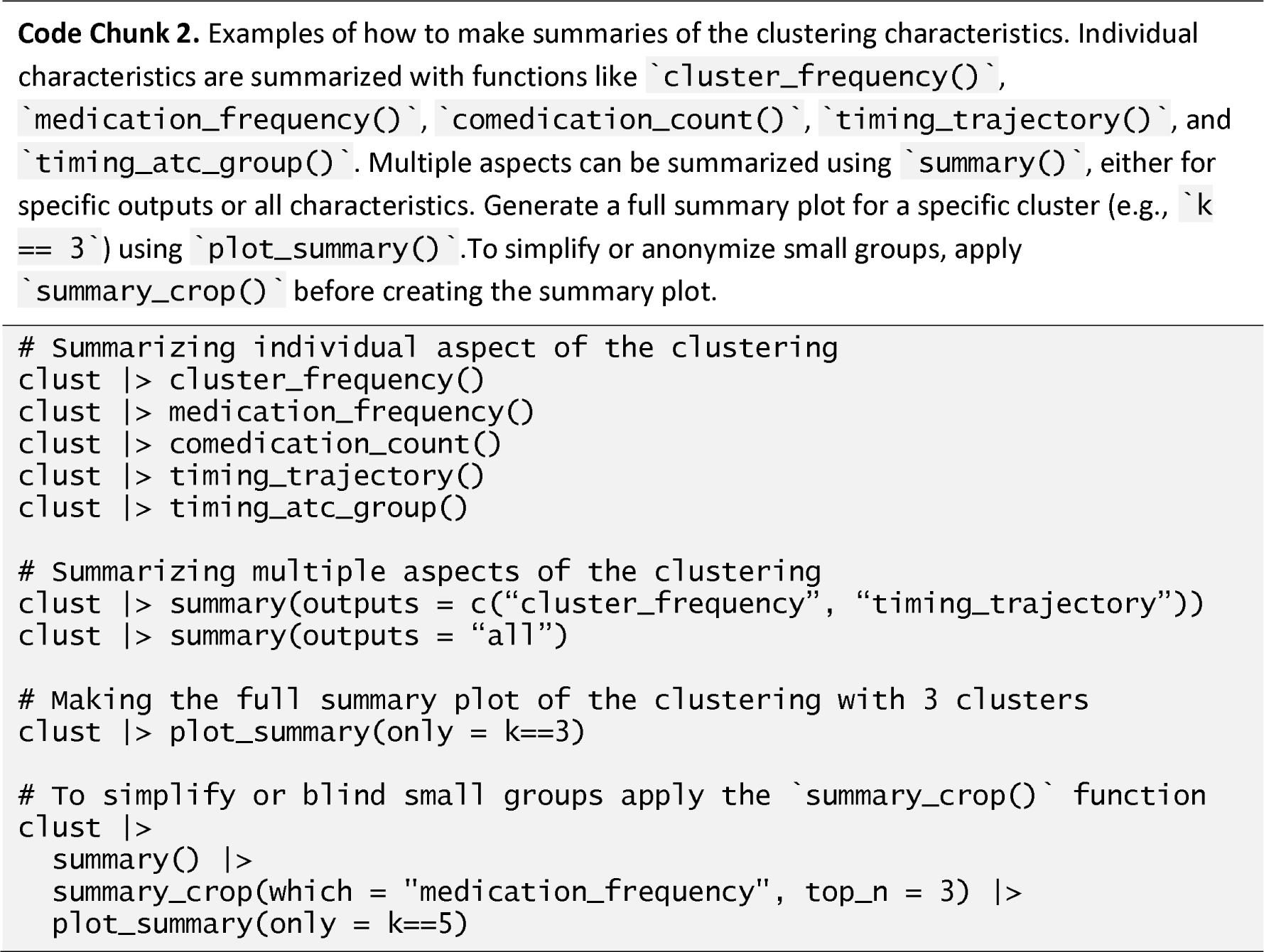

**Figure 2.**
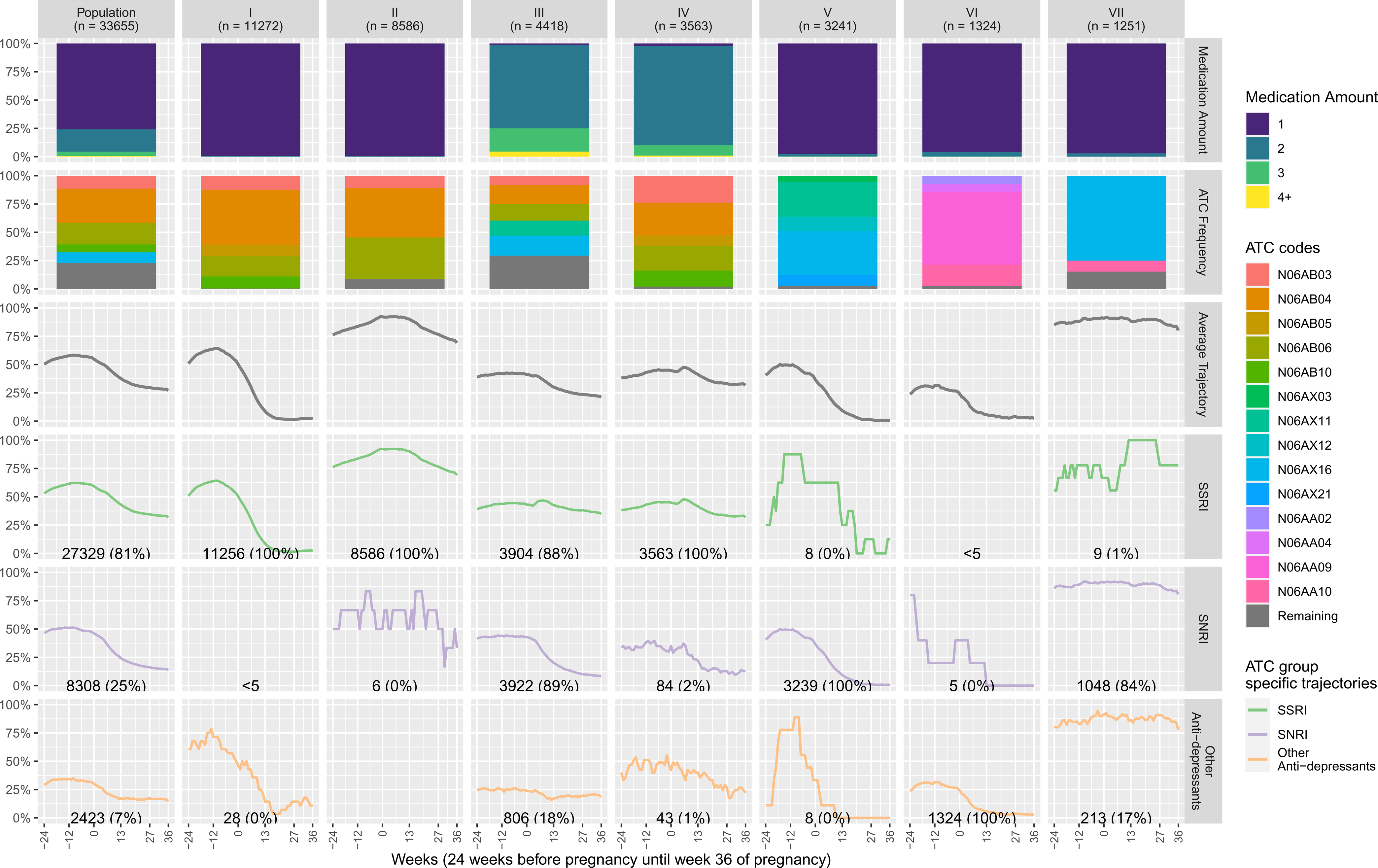
Characteristics of medication use clusters in a cohort of 33,655 Danish pregnant women with at least one antidepressant prescription within the six months before pregnancy. The figure illustrates the clustering of these medication patterns, learned from the data using the ‘medic()’ function and visualized with ‘plot_summary()’. The first column represents the entire population, while the subsequent seven columns depict the characteristics of each cluster, revealing distinct patterns. Each row provides a different characteristic: the first row illustrates the number of antidepressants used, the second row illustrates the 5 most prevalent antidepressant ATC codes, and the remaining 4 rows illustrate the average antidepressant exposure timing by anti-depressive type.

‘employ()’: Applying Clusters to New Data

An important functionality of the package is the ‘employ()’ function, which takes an existing clustering and a new dataset and uses this particular clustering on the new data. Each new individual is assigned to the closest existing cluster. This functionality enables the learning of the clustering on a sub-cohort such that this clustering can be applied to the entire cohort. This is especially advantageous for saving computational time by clustering on a representative sub-cohort and then generalizing, or when working with a distinct sub-cohort, which should guide the learning of medication features (e.g., persons with a specific diagnosis or persons exposed to a specific interest medication). Moreover, this functionality enables easy sharing of learned clusters across studies and countries.

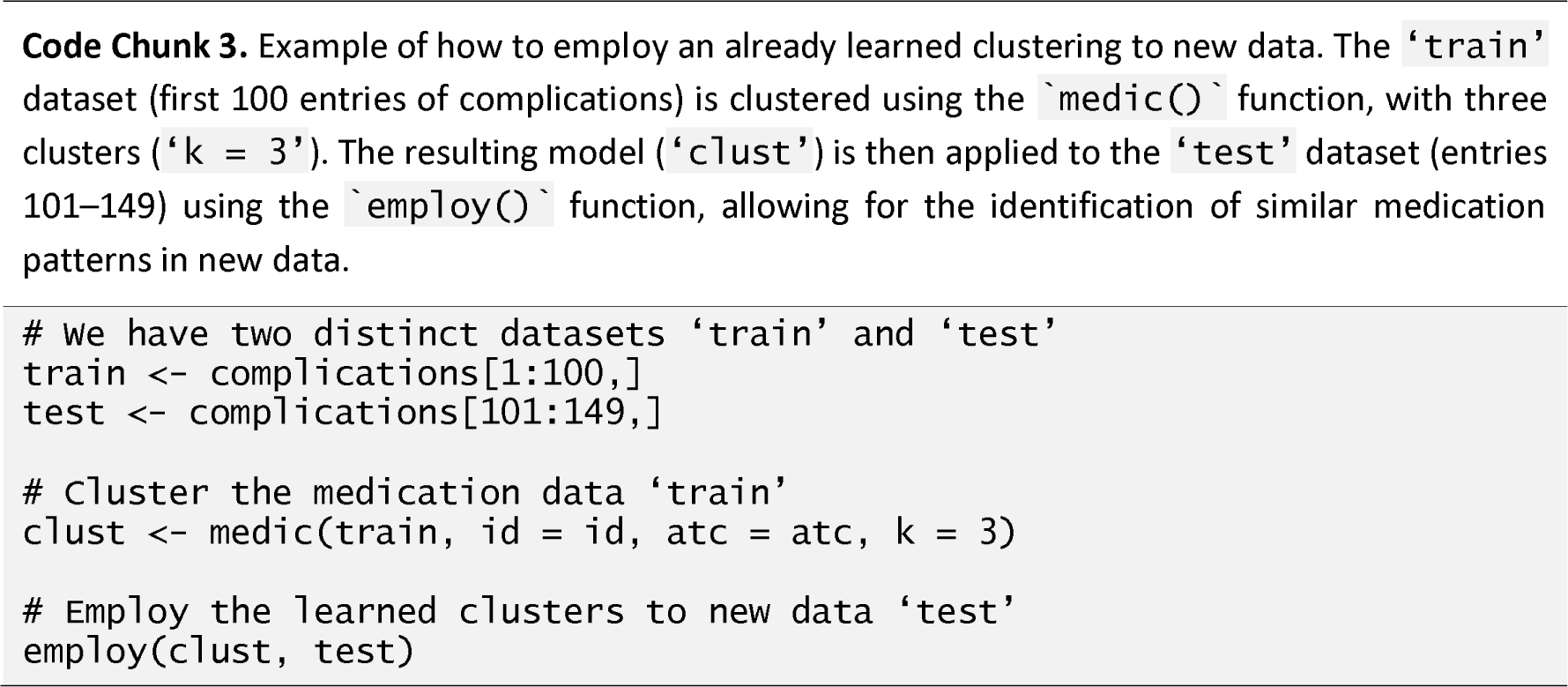

### Installation

The tame package is available on The Comprehensive R Archive Network (CRAN), and may be installed by running ‘install.packages(“tame”)’ in R ≥4.2. Additionally, development versions may be available on github at https://github.com/Laksafoss/tame.

## USE

To demonstrate the use and utility of tame, we apply the methodology to the medication use immediately before- and during pregnancy using a cohort of Danish women with a history of antidepressant use before pregnancy. We investigate how these medications are used in practice, and how use is related to redeeming psycholeptics in the first year after pregnancy. Please note that this study primarily serves as an example of how to use the package.

### Ethics declaration

The Danish national registries used in this study are protected by the Danish Data Protection Act. No informed consent nor approval from the Danish Research Ethics Committees were needed for this study, since only national register data was used. By law, no ethics committee approval is required for studies only using register-data.

### Characterizing Antidepressant Use Before and During Pregnancy

We identified all live-born singleton births with a gestational age of at least 36 weeks between 1997 and 2016 in the Danish Medical Birth Registry [10]. Using the Danish National Prescription Registry, the cohort was restricted to women who redeemed at least one antidepressant within the six months before pregnancy. To conduct outcome analyses we also linked this cohort to a number of registers on demographic, socioeconomic and healthcare information; for more on these variables see S6. The anonymized individual-level data was accessed on Denmark Statistics’ researcher machines on December 12th, 2023.

Assuming an exposure of WHO’s defined daily dose each day, we estimated the time varying antidepressant exposure of each individual from 24 weeks before pregnancy until gestational week 36 in the following medication groups: selective serotonin reuptake inhibitors (SSRI; ATC codes starting with N06AB), serotonin-norepinephrine reuptake inhibitors (SNRI; ATC codes starting with N06AX), and others antidepressants (ATC codes starting with N06AA, N06AF or N06AG). For each individual and each day, antidepressant use was classified as exposed or unexposed (0/1). A description and discussion of the tuning parameters chosen for this example may be found in S3. Seven clusters were used to describe the data.

Figure 2 characterizes the clustering of antidepressant medication patterns learned from the data using ‘medic()’ and then illustrated with ‘plot_summary()’. In the first column, the characteristics of the entire population are illustrated, and the following 7 columns display the characteristics of each cluster, highlighting distinct patterns within the data. Each row illustrates a different characteristic: number of antidepressants used per person in the cluster, frequency of use of the 5 most prevalent antidepressant ATC codes in the cluster, average cluster timing of all antidepressant medications, SSRI use, SNRI use, other antidepressant use, respectively.

For the entire study population (column 1), row 1 demonstrates that the majority (76%) of the mothers take only one antidepressant in the study period, one-fifth are exposed to two antidepressants, and even fewer (4%) are exposed to three or more antidepressants. In row 2, we see that four out of five of the most used antidepressants are SSRIs. Rows 3 to 6 show the average medication exposure trajectory among all medications (row 3), among SSRIs (row 4), among SNRIs (row 5), and among other antidepressants (row 6). As we are considering a binary exposure in this example, the average displayed may be interpreted at the percentage of pregnancies exposed at that particular timepoint. The overall average (row 3) shows that at any given week before the conception, at least half of the studied women are exposed to one or more antidepressants; then, during the 12 first weeks of pregnancy, the antidepressant use drops. This pattern is consistent across all three groups of antidepressants, but the level of use in any given week is different, with SSRI usage being consistently more frequent than SNRI and other antidepressants (rows 3-6). We further describe clusters II, III, and IV here, and all clusters in S8.

Cluster II – “Sustained use of one SSRI”: The second cluster, with 26% of pregnancies, is almost exclusively SSRI single drug users. The pregnancies in this cluster has a high level of sustained SSRI usage with at least 70% of pregnancies being exposed at any time point in the study period, and with more than 90% of pregnancies being exposed in the weeks 0 to 13.

Cluster III – “Concomitant use of different types of antidepressants”: The third largest cluster with 13% of pregnancies studied, is characterized by concomitant drug use of a mix of all 3 classes of antidepressants. More than 88% of pregnancies are exposed to either SSRI or SNRI or both, while only 18% of pregnancies are exposed to other antidepressants. The SSRI use frequency remains fairly stable across the entire exposure period, as opposed to the SNRI use frequency which drops at the start of the pregnancy.

Cluster IV – “Multiple SSRIs used”: Cluster IV, which consists of 11% of pregnancies is characterized by multiple drug use, with less than 3% using only one drug, almost 88% using 2 antidepressants, and 10% using 3 or more antidepressants. Unlike cluster III, where pregnancies are exposed to a mix of different types of antidepressants, in this cluster, the exposure is multiple SSRIs.

### Risk of redeeming psycholeptics within one year of birth according to medication cluster membership

To give an example of how to use these learned medication clusters in further analysis, we apply them in an analysis of the adjusted cumulative incidence of adverse psychiatric post-partum outcomes. In this example, redemption of psycholeptics (ATC group N05; anti-psychotics N05A, anxiolytics N05B, and hypnotics and sedatives N05C) in the first year following birth is considered as the outcome, as it serves as a concrete indicator of psychiatric distress and healthcare utilization, directly impacting maternal and child well-being.

We used stabilised inverse probability of treatment weighting. Weights were calculated as the inverse odds of being assigned medication cluster I, III, IV, V, VI or VII versus medication cluster II adjusted for confounders (see S8 for the full list) in a multinomial regression model. Cluster II was chosen as the reference cluster as it is a large cluster (26% or pregnancies) with a simple interpretation (sustained use of one SSRI throughout the exposure period).

Risk ratios and risk differences were then calculated at the 365-day mark. The results of the adjusted cumulative incidence analysis are shown in Figure 3a, and the estimated risk ratio and risk difference by medication cluster are shown in Figure 3b.

**Figure 3.**
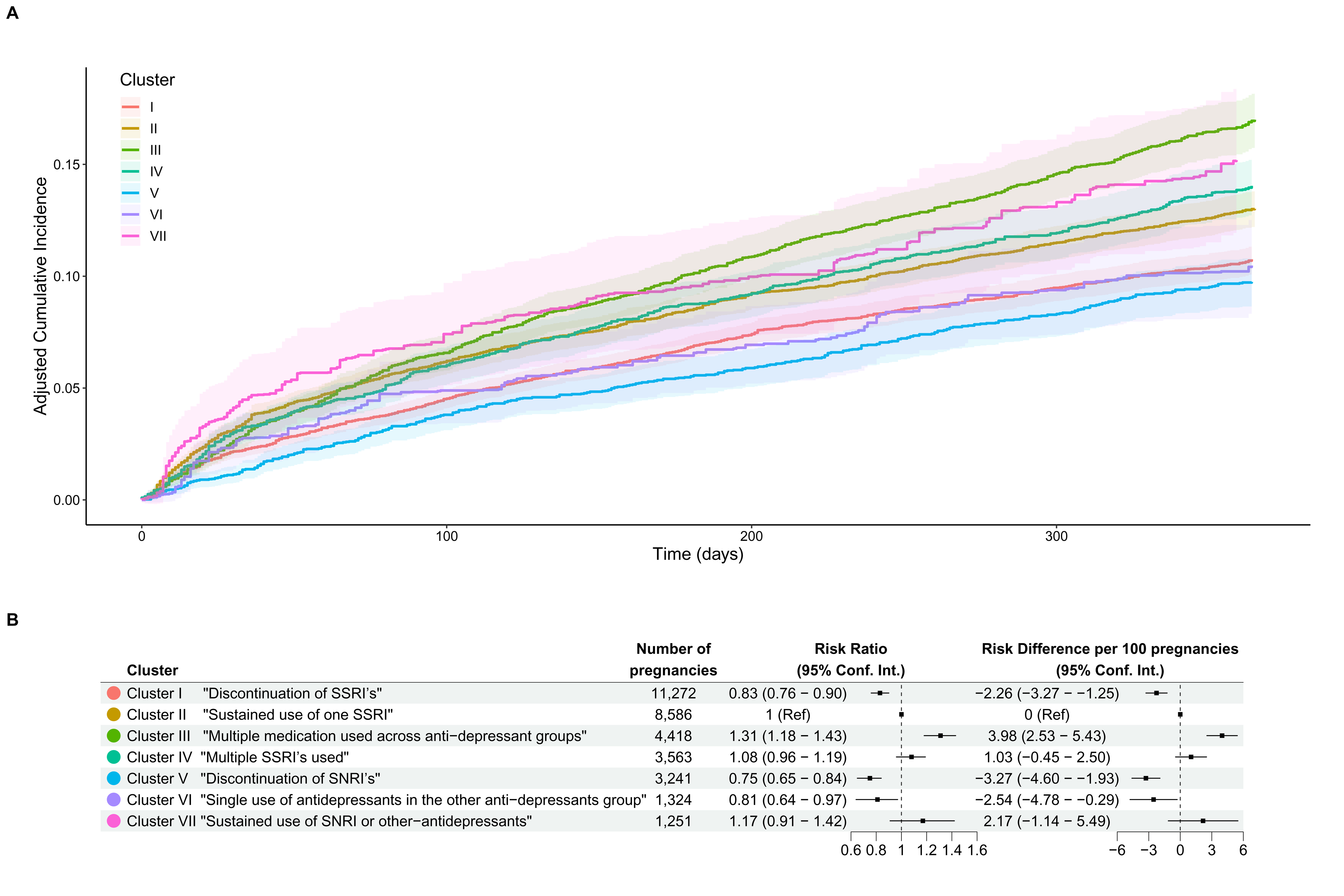
Adjusted* risk of redeeming psycholeptics in the first year following birth according to medication cluster membership. (A) Adjusted cumulative incidence of redeeming psycholeptics in the first year following birth according to medication cluster membership. (B) Adjusted risk ratio and risk difference per 100 pregnancies of redeeming psycholeptics one year after birth according to medication cluster membership. * Both absolute (A) and relative (B) risks were adjusted for maternal age (<25, 25-29, 30-34, ≥35), parity (0, 1, 2, ≥3), BMI (<18.5, 18.5-25, 25-30, 30-35, >35), smoking status (non-smoker, stopped smoking or smoker), family structure (married, single, or living with partner), maternal employment status (employed, employed in a management position, self-employed, or unemployed and receiving public assistance), maternal level of education (primary, secondary, postsecondary, or vocational school), location of residence in Denmark (capital region, central region, northern region, Zealand, or southern region), disposable household income (quartile 1, 2, 3, or 4), and maternal country of origin (Denmark, Europe (without Denmark), or other), history of psychiatric hospitalizations, history of self-harm and Charlson comordibity score ≥1.

From Figure 3b we see that three clusters have a significantly reduced risk of redeeming psycholeptic prescriptions within 1 year of birth as compared with cluster II, sustained exposure to one SSRI. These three clusters are cluster I (“discontinuation of SSRI’s”), cluster V (“discontinuation of SNRI’s”) and cluster VI (“single use of antidepressants in the other antidepressants group”). Conversely, cluster III (“multiple medication used across antidepressant groups”) is significantly associated with an increased risk of redeeming psycholeptics within 1 year of birth as compared with cluster II (“sustained use of on SSRI”).

We can also observe from Figure 3b that neither cluster IV (“multiple SSRI’s used”), nor cluster VII (“sustained use of SNRI or other-antidepressants”) where significantly associated with a higher risk of redeeming psycholeptics as compared with cluster II (“sustained use of on SSRI”). All statistical conclusions were supported on both the risk ratio and risk difference scale.

## DISCUSSION

The tame package implements a novel data-driven learning method for understanding individual-level medication patterns through hierarchical clustering and provides tools for illustrating and characterising these clusters. The construction of a distance measure is the cornerstone of any clustering method, as it dictates the method’s ability to accurately group similar observations and differentiate dissimilar ones. In machine learning, the quality of a distance measure directly impacts the success of the model. Our novel distance measure, specifically tailored for medication data, uniquely incorporates the ATC code, exposure timing, and medication dosage, allowing researchers to fine-tune the relative importance of these factors. This flexibility is crucial for ensuring that the clusters reflect the true nature of the data and the research objectives. Moreover, a method for applying existing medication clusters to new medication pattern data has been developed.

Here, we demonstrated how tame can help identify and narrow down complex trends of antidepressant use before- and during pregnancy, using a national Danish cohort. In general, clustering real-world medication data offers a valuable automated search method for detecting potential safety signals or adverse drug interactions that may not have been previously suspected. Unlike traditional approaches that rely on specific suspicions or hypotheses, clustering allows for an exploratory analysis of medication patterns. For example, through clustering, researchers may discover that a particular cluster, such as medications X and Y frequently taken together, is associated with an elevated risk of adverse outcomes. This automated identification of medications and medication combinations that pose potential risks can serve as an early warning system, prompting further investigation into these specific medication combinations and their effects.

Moreover, real-world medication patterns captured through clustering analysis reflect the diverse medication usage habits of individuals outside controlled clinical trial settings. This real-world complexity adds depth to our understanding of medication usage and its implications. Additionally, these patterns can be leveraged for statistical adjustment or stratification in epidemiological studies. By accounting for these real-world medication patterns, researchers can better control for confounding factors and obtain more accurate estimates of treatment effects in epidemiological studies and observational research.

When using this method, specifying tuning parameters provides great customizability but require decisions from the researcher. Thus, tuning the distance measure will still have to be done in accordance with the understanding of the studied medication and the clinical setting. However, methods for algorithmically optimizing the number of clusters and assisting in the choice of linkage according to measures of goodness of fit is currently underway.

Naturally, clustering personal medication usage according to ATC codes is limited by the ATC classification system itself. The ATC hierarchy classify according to main therapeutic use or mechanism of action of the main active ingredient, and as such, encode the main indication of the drug. However, many medications are used for multiple indications but will only be assigned one ATC code according to the main indication. This can obscure the full spectrum of a medication’s applications. Moreover, an ATC group may be specified according to mechanism of action, resulting in groups where the medications have diverse indications, complicating the interpretation of clustering results. In addition, a chemical substance may be given more than one ATC code if it is used for two different therapeutic purposes. For a detailed introduction to the construction of WHO’s ATC classification system see [11]. Thus, as the ATC classification system classifies both according to the therapeutic and the pharmacological aspects of a medication, so does the classification method presented in this paper. Methods for extending the ATC distance measure to allow for user defined exceptions or additions to the ATC classification system are in progress. This extension will allow the user to give all medications with identical active ingredients a smaller distance regardless of distance in the ATC hierarchy or make combination products more similar than the ATC hierarchy suggests.

In addition, a central feature of hierarchical clustering, and many other clustering methods, is that all observations are assigned a cluster. Thus, if the data contain many outliers, which in fact are not comparable with other observation in the data, these will still be placed in clusters. Moreover, the assignment to a cluster is done in a deterministic way, and the method does not provide uncertainty estimates for this assignment. These features of clustering lead to less robustness to changes to the study population. When using tame, the ‘employ()’ function can be used to gain some insight into the extent of this problem in a given dataset through cross validation type strategies.

Lastly, it should be noted that clustering methods can be very computationally time and RAM demanding. This means that we may still be limited in our applications of this method to large cohorts with wide medication use by our computational power. As discussed in S4, this central limitation has informed multiple design aspects of the code itself, and computationally intensive parts of the code are written in C++ or utilize code written in Fortran. This works to ensure a relatively respectable computational time for our example with just over 33,000 antidepressant exposed pregnancies of less than 10 minutes on a system with Intel(R) Xeon(R) Gold 6254 CPU @ 3.10GHz.

## Conclusions

In summary, the tame package improves on classical approaches to understanding drug exposure by clustering complex, real-world use patterns which integrate information on timing, dose, and type of medication. Here, we described an application of how we have used this method to understand antidepressant use up to and during pregnancy, and then analyse how these learned clusters were associated with the use of psycholeptics in the first year following birth. In the future, the method may be used to understand other patterns of prescription drug use and optimize drug safety surveillance through data driven learning.

## ACKNOWLEDGMENTS

Authors thank Kim Daniel Jakobsen for his invaluable assistance in the development of the methodology employed in this study. We would also like to thank Elisabeth O’Regan for her meticulous attention to detail in formatting and refining the English language of this manuscript.

## AUTHOR CONTRIBUTIONS

Conceptualising R package functionalities: A.D.L. and J.W. Code development and documentation: A.D.L. Code testing: A.D.L. Study concept and design: A.D.L., J.W. and A.H.. Data analysis: A.D.L. Manuscript writing: A.D.L. Critical review: A.D.L., A.H. and J.W. Funding: A.H.

## DATA AVAILABILITY STATEMENT

Data cannot be shared publicly because of data privacy regulations in Denmark. Applications for data access should be submitted to Research Services at the Danish Health Data Authority.

## COMPETING INTERESTS STATMENT

The authors declare no competing interests.

## REFERENCES

[1] C. Hurault-Delarue, C. Chouquet, N. Savy, I. Lacroix, A.-B. Beau, J.-L. Montastruc and C. Damase-Michel, “How to take into account exposure to drugs over time in pharmacoepidemiology studies of pregnant women?,” Pharmacoepidemiology and Drug Safety, vol. 25, no. 7, pp. 770–777, 2016.

[2] M. E. Wood, A. Lupattelli, K. Palmsten, G. Bandoli, C. Hurault-Delarue, C. Damase-Michel, C. D. Chambers, H. M. E. Nordeng and M. M. H. J. van Gelder, “Longitudinal Methods for Modeling Exposures in Pharmacoepidemiologic Studies in Pregnancy,” Epidemiologic Reviews, vol. 43, no. 1, pp. 130–146, 2021.

[3] L. S. Lemon, L. M. Bodnar, W. Garrard, R. Venkataramanan, R. W. Platt, O. C. Marroquin and S. N. Caritis, “Ondansetron use in the first trimester of pregnancy and the risk of neonatal ventricular septal defect,” International Journal of Epidemiology, vol. 49, no. 2, pp. 648–656, 2019.

[4] D. Steinley, “K-means clustering: A half-century synthesis,” British Journal of Mathematical and Statistical Psychology, vol. 59, no. 1, pp. 1–34, 2006.

[5] C. Genolini, X. Alacoque, M. Sentenac and C. Arnaud, “kml and kml3d: R Packages to Cluster Longitudinal Data,” Journal of Statistical Software, vol. 65, no. 4, pp. 1–34, 2015.

[6] C. Genolini, B. Falissard and P. Kiener, “R package version 2.4.6,” 2023. [Online]. Available: https://CRAN.R-project.org/package=kml.

[7] C. Proust-Lima, V. Philipps and B. Liquet, “Estimation of Extended Mixed Models Using Latent Classes and Latent Processes: The R Package lcmm,” Journal of Statistical Software, vol. 78, no. 2, p. 1–56, 2017.

[8] S. Salvatore, D. Domanska, M. Wood, H. Nordeng and S. Geir Kjetil, “Complex patterns of concomitant medication use: A study among Norwegian women using paracetamol during pregnancy,” PLOS ONE, vol. 12, no. 12, pp. 1–11, 2017.

[9] A. Laksafoss, tame: Timing, Anatomical, Therapeutic and Chemical Based Medication Clustering, 2023-02-23.

[10] M. Bliddal, A. Broe, A. Pottegård, J. Olsen og J. Langhoff-Roos, »The Danish Medical Birth Register,« European Journal of Epidemiology, årg. 33, nr. 1, pp. 27-36, January 2018.

[11] Health, Norwegian Institute of Public, Last updated: 2022-11-10. [Online]. Available: https://www.whocc.no/atc/structure_and_principles/. [Senest hentet eller vist den 2023-12- 19]].

[12] N. T. H. Trinh, T. Munk-Olsen, N. R. Wray, V. Bergink, H. M. E. Nordeng, A. Lupattelli and X. Liu, “Timing of Antidepressant Discontinuation During Pregnancy and Postpartum Psychiatric Outcomes in Denmark and Norway,” JAMA Psychiatry, Published online March 08, 2023.

[13] A. Pottegård, S. A. J. Schmidt, H. Wallach-Kildemoes, H. T. Sørensen, J. Hallas and M. Schmidt, “Data Resource Profile: The Danish National Prescription Registry,” International Journal of Epidemiology, vol. 46, no. 3, pp. 798–798f, 2017.

[14] M. Bliddal, A. Broe, A. Pottegård, J. Olsen and J. Langhoff-Roos, “The Danish Medical Birth Register,” European Journal of Epidemiology, vol. 33, no. 1, pp. 27–36, January 2018.

