## Supplementary Material for "tame: An R package for identifying clusters of medication use based on dose, timing and type of medication"

### Supplementary 1. Background Mathematics

#### The anatomical therapeutic chemical classification system

The Anatomical Therapeutic Chemical (ATC) Classification System is a drug classification system developed by the World Health Organization (WHO) to assist in drug monitoring and research. In this drug classification system, the active ingredients are classified using a hierarchy with five levels.

At the first level – the main level – there are 14 distinct groups based on anatomical and pharmacological differences in drug use. This group is indicated by a letter, e.g. "A" for Alimentary Tract and Metabolism or "J" for Anti-infectives for Systemic Use. Each of these 14 main groups then branch off into increasingly subdivided groupings of individual drugs according to therapeutic, pharmacological and chemical subgroups, and finally into chemical substances. At each level, new symbols are added to the ATC code; the second level adds two digits, the third and fourth level each append a letter, and finally the fifth level adds two digits.

As a result of this strict hierarchy of encoding, one can always derive all lower level codes from any ATC code. We let $L^{v}$ denote the function that extracts the level $v$ code from an ATC code, so that e.g. $L^{1}\left( N06AB04 \right)=N$ and $L^{3}\left( B03BA \right)=B03B$.

#### Medication profiles and patterns

We consider individual level *medication profiles* $p_{i}$ for each individual $i=1,\ldots, n$. An individual’s medication profile,

$$p_{i}=\left( q_{im} \right)_{m\in M_{i}} ,$$

(1)

consists of *medication patterns* $q_{ik}$ which encode personal and medication specific information for each *medication* that person was exposed to ${m\in M}_{i}$,

$$q_{im}=\left( a_{im}, \left( d_{imt} \right)_{t\in T} \right).$$

(2)

Here, $a_{im}$ denotes the ATC code of individual $i$’s medication $m$, and $d_{imt}\in D$ is the dose intensity at timepoint $t$ on the shared timescale $T$ of said medication. The choice of D and T will be governed by the research question. In supplementary 5 below a number of example choices are illustrated.

#### Distance measure

Using this notation, we introduce our two central distance measures. These two measures provide different strategies for measuring the difference between two individual level medication profiles.

The first measure, *the average distance measure*, simply computes the average distance of all comparisons between two people’s medication patterns giving each comparison equal weight

$$d^{a}\left( p_{i},p_{j} \right)=\frac{1}{\left( \left| M_{i} \right|\left| M_{j} \right| \right)^{\alpha}}\sum_{m\in M_{i}} \sum_{n\in M_{j}} C\left( q_{im}, q_{jn} \right),$$

(3)

while the second measure, *the similar representatives distance measure*, computes the average of the one-sided comparisons of the most similar representatives

$$d^{s}\left( p_{i},p_{j} \right)=\frac{1}{2\left| M_{i} \right|^{\alpha}}\sum_{m\in M_{i}} \min_{n\in M_{j}} C\left( q_{im},q_{jn} \right)+\frac{1}{2\left| M_{j} \right|^{\alpha}}\sum_{n\in M_{j}} \min_{m\in M_{i}} C\left( q_{im},q_{jn} \right).$$

(4)

Both measures rely on the more basic measure, *the comparison measure*, which measures the distance between two medication patterns

$$C\left( q_{im}, q_{jn} \right)=\left( \left( 1+W\left( a_{im},a_{jn} \right) \right)\left( 1+\gamma S\left( \left( d_{imt} \right)_{t\in T},\left( d_{jnt} \right)_{t\in T} \right) \right)-1 \right)^{\beta} ,$$

(5)

where the *ATC distance measure*, $W$, is defined as

$$W\left( a_{im},a_{jn} \right)=\theta_{l} where l=\underset{v\in\{0, .., 5\}}{\mathrm{argmax}} L^{v}\left( a_{im} \right)=L^{v}\left( a_{jn} \right)$$

(6)

and the *dose trajectory distance measure,* $S$*,* is given as the Minkowski distance between the two dose trajectories

$$S\left( \left( d_{imt} \right)_{t\in T},\left( d_{jnt} \right)_{t\in T} \right)=\left( \sum_{t\in T} \left| d_{imt}-d_{jnt} \right|^{p} \right)^{1/p}.$$

(7)

#### Hierarchical clustering and linkage criteria

The medic() function implements an agglomerative hierarchical clustering scheme. This analysis constructs a cluster hierarchy by initially treating each observation as its own cluster. Subsequently, clusters are iteratively merged based on similarity, ultimately forming a hierarchical structure. The merging process continues until the desired number of clusters is achieved. To decide which pair of clusters to merge, the distance between clusters is considered. This cluster similarity is determined by a linkage criterion. The distance measures defined above specifies the distance between any pair of observations, and the linkage criterion specifies the distance between sets of observations.

The following linkage criteria are supported in medic().

| **Supplementary Table 1.** Overview of the linkage criteria between two sets of observations $A$ and $B$ implemented in medic(). | | |
| --- | --- | --- |
| **Criterion Name** | **Formula** | **The method name in R** |
| Complete linkage | $\max\left\{ d\left( a,b \right):a\in\left\vert A \right\vert, b\in\vert B\vert\right\}$ | “complete” |
| Single linkage | $\min\left\{ d\left( a,b \right):a\in\left\vert A \right\vert, b\in\vert B\vert\right\}$ | “single” |
| Average linkage | $\frac{1}{\left\vert A \right\vert\cdot\vert B\vert}\sum_{a\in\vert A\vert} \sum_{b\in\vert B\vert} d\left( a,b \right)$ | “average” |
| Ward’s linkage | $ESS\left( A\cup B \right)-\left( ESS\left( A \right)+ESS\left( B \right) \right),$  Where $ESS\left( A \right)=\sum_{a\in\vert A\vert} \left\vert a-\frac{1}{\vert A\vert}\sum_{a\in\vert A\vert} a \right\vert$ | “ward.D2” |

### Supplementary 2. Mathematical Glossary

| *DATA ENCODING* | | |
| --- | --- | --- |
| $p_{i}=\left( q_{im} \right)_{m\in M_{i}}$ | Medication profile | Individual $i$’s complete medication information consisting of a number of personal medication patterns. |
| $q_{i}=\left( a_{im}, \left( d_{imt} \right)_{t\in T} \right)$ | Medication pattern | Individual $i$’s information on a specific medication |
| $M_{i}$ | Medication | The complete list of medications individual $i$ was exposed to. |
| $a_{im}$ | ATC code | The WHO ATC code of the $m$’th medication of individual $i$. E.g. A05BB05 |
| $d_{imt}$ | Dose intensity | The dose at time $t$ of the person specific medication. E.g. 0/1 encoding exposed yes/no, or a positive number indication the percentage of the DDD. |
| $T\mathbb{\subseteq N}$ | Timescale | The shared timescale. This timescale must be shared and consistent across all study participants. E.g. weeks from conception to week 36 of a full-term pregnancy, or days since hospitalization. |
| $D\mathbb{\subseteq R}$ | Dose Space | The shared space of possible doses. The dose space must be consistent across all medication and study participants. E.g. exposed yes/no, or a percentage of defined daily dose. |
| *DISTANCE MEASURE* | | |
| $d^{a}$ | Average measure | |
| $d^{s}$ | Similar representatives measure | |
| $C$ | Pattern comparison measure | |
| $W$ | ATC distance measure | |
| $S$ | Dose trajectory distance measure | |
| $\alpha$ | Normalization tuner | |
| $\beta$ | Dis-similarity penalizing tuner | |
| $\gamma$ | Timing importance tuner | |
| $p$ | Minkowski distance parameter | |
| *CLUSTERING* | | |
| $k$ | Cluster index | The index of a clusters $C_{k}$ in a constellation $C$ |
| $C_{k}$ | A cluster | Cluster $k$ in the constellation $C$ |
| $C=\left( C_{k} \right)$ | A constellation | The collection of all the clusters resulting from a run of the algorithm. |

### Supplementary 3. Tuning the method and interpreting parameters

#### Choosing number of clusters

Choosing the number of clusters is always one of the central challenges of hierarchical clustering, and it can be hard to give general advice. As this method is intended to be used within the fields of epidemiology, medicine and health sciences, we advise that clinical interpretability is the primary guide when choosing the number of clusters rather than relying solely on algorithmic methods such as elbow plots.

Algorithmic methods for determining the number of clusters in hierarchical clustering often overlook qualitative aspects, potentially leading to suboptimal choices, especially with heterogeneous and high-dimensional data like medication usage records. Traditional methods may fail to provide clear guidance in such cases. Instead, researchers should align the number of clusters with their specific clinical questions. For instance, a lower number may suffice for studies seeking to characterizing usage patterns, while a higher number may be needed for safety signal detection or subgroup exploration.

A suggested approach is to define a narrow range of acceptable cluster numbers, such as 8-10, and then apply algorithmic methods for assistance in making the final decision. However, visual inspection of clusters remains essential for assessing clinical relevance and interpretability.

#### Choice of linkage criteria

The linkage is the measure of dissimilarity between sets of observations. This measure is used to decide which clusters to combine. Thus, the choice of linkage can have a large impact on the resulting constellation. The complete linkage criterion tends to find compact clusters of approximately equal diameters, while the single linkage, also known as the friends of friends method, tends to produce thin and long clusters. All other linkage methods can be viewed as somewhere between these two methods. As the average and complete linkages tend to yield more balanced dendrograms, these linkages are often preferred.

#### Choice of distance measure

To find the distance between two persons’ medication profiles, our measure summarizes the distances between the individual medication patterns. We have implemented two general summary methods, namely, the “average” method and the “similar representatives” method.

The “average” method reflects the belief that all aspects of a person’s medication pattern are equally relevant at all times, while the “similar representatives” method supposes that for each comparison, only one medication pattern is necessary for understanding the relation to another person’s medication pattern. Therefore, the “average” method may be more suitable when the medication data being analyzed are narrow and more homogeneous - that is, only a smaller subset of closely related medication types are investigated. Conversely, the “similar representatives” method tends to be well-suited for wide and heterogeneous medication data, such as investigations of all medication exposures over a given period.

#### Tuning parameter for the distance measure

##### Theta

The six theta parameters, $\theta=\left( {\theta_{0},\theta}_{1}, \theta_{2}, \theta_{3}, \theta_{4},\theta_{5} \right)$, encode the relative importance of each ATC level among the medications under study. The value $\theta_{0}$ encodes the maximum distance between two ATC codes. This is the distance assigned when the ATC codes only match on the zeroth level, that is, there is no comparison between the codes, i.e. $W\left( N06AB02,B03AA01 \right)=\theta_{0}$. The ATC distance $\theta_{1}$ is assigned when ATC codes match on level 1 and no deeper, and so on for each level.

In this way, we can assert assumption on the relative importance of the ATC levels appropriate for the medication under study. If the exact chemical compound is essential for understanding the difference between the chosen medications, then a $\theta$ that has a large difference between $\theta_{5}$ (difference on level 5) and zero (identical ATC codes) and smaller differences between $\theta_{4}$, $\theta_{3}$,$\theta_{2}$,$\theta_{1}$ and$\theta_{0}$ is appropriate. Examples of this can be seen illustrated in supplementary figures 1A, 1B and 1G below, where the example $\theta$ illustrated in 1G is the most extreme: assuming that only the first level of the ATC code is of importance.

If the anatomical or pharmacological aspects of a drug is more important for understanding the differences between the drugs under study, then a $\theta$ with large gaps between $\theta_{0}$, $\theta_{1}$ and $\theta_{2}$, and comparatively smaller gaps between $\theta_{3}$, $\theta_{4}$, $\theta_{5}$ and zero (identical ATC codes) is appropriate. Examples of this is illustrated in supplementary figures 1E, 1F, 1K and 1L, with figures 1K and 1L illustrating the extreme case where only level 2 and level 1 are of importance.

If one does not have any strong assumptions on relative ATC level importance choosing a moderate $\theta$ where each level is given equal importance, like the example illustrated in figure 1D, may be a good choice.

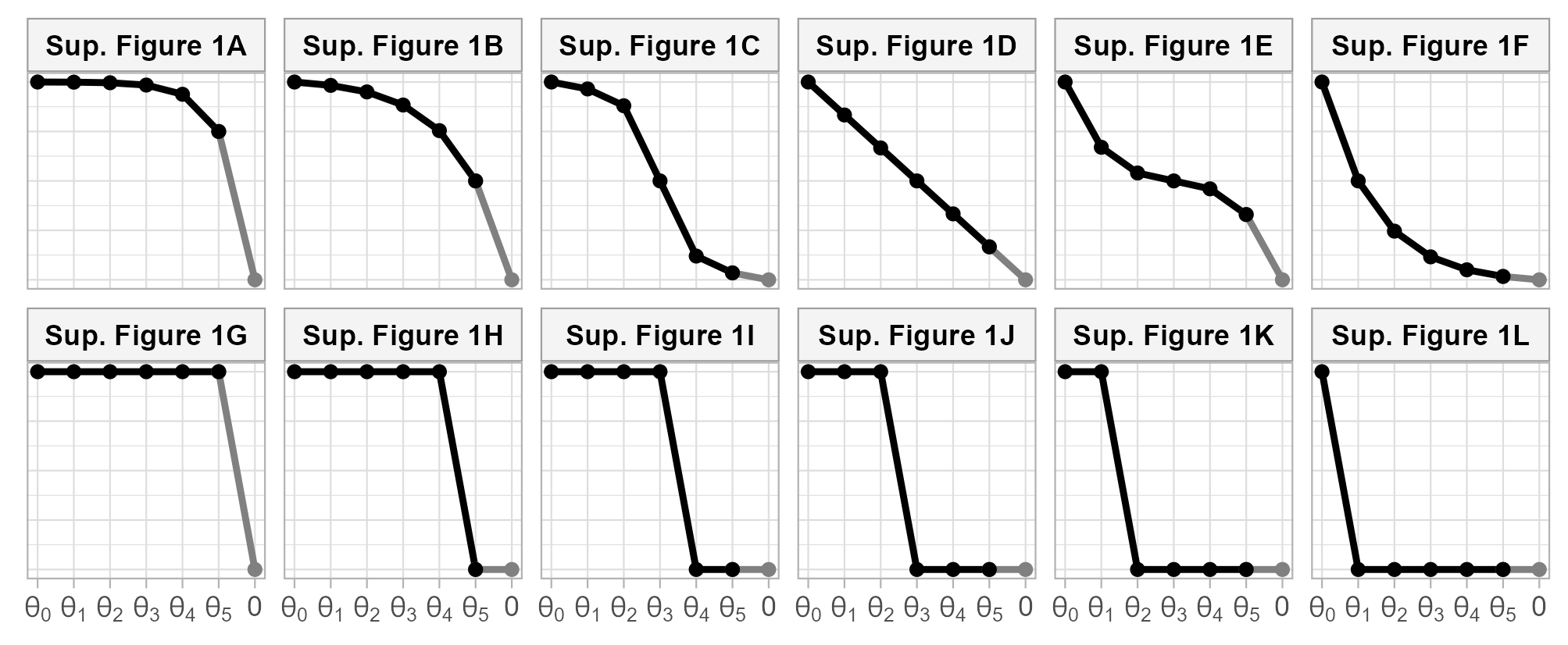

**Supplementary Figure 1.** Example choices of relative values of $\theta_{0}$, $\theta_{1}$, $\theta_{2}$, $\theta_{3}$, $\theta_{4}$ and $\theta_{5}$ emphasizing different relative importance of the difference ATC levels.

##### Gamma

The gamma, $\gamma$, parameter tunes the relative importance of the dose-timing aspect and the ATC of a medication pattern. A larger $\gamma$ gives more weight to the timing aspect. General guidelines for tuning $\gamma$ is harder to make, as it must always be tuned in relation to $\theta$ and the range of the dose intensities observed.

Choosing $\gamma$ such that

$$\gamma\cdot\max_{i,m,t} d_{imt}=\theta_{0},$$

(1)

weights the dose intensity trajectory and the ATC codes equally, while choosing a larger $\gamma$ emphasizes the dose intensity more, and conversely choosing a smaller $\gamma$ assumes that the ATC codes ought to hold more weight.

### p

The dose trajectory distance measure is simply the Minkowski distance on the dose intensity trajectories and is tuned in the usual way with $p\geq1$. Typically, one uses $p$ equal to 1 or 2, corresponding to the Manhattan and Euclidean distance, respectively. The larger the $p$ chosen, the more extreme distances between dose intensity trajectories is penalized.

When considering medications where small changes in dosage or timing is of large importance it is usually suitable to choose a larger $p>1$. Whereas, medications with a wide range of dosages used may benefit from a lower $p<2$. Choosing a smaller $p$ may also be preferable when there is uncertainty surrounding the exact dosage or timing, so as to not penalize recoding errors to harshly.

Note that as the number of time observations increase it becomes preferable to choose a smaller $p$, as nearness in higher dimensions becomes a less well-defined concept (1). A general conservative recommendation, is to choose $p=1$ to maintain our intuition of proximity as dimensions rise.

##### Alpha

Both summary methods involve normalization over the number of medications. The strength of this normalization is tuned through $\alpha>0$. Using $\alpha=1$, the default, gives averages across all the comparisons made in each summary method. This ensures that people who take a lot of medications do not become outliers. However, if one expects that people who take a larger number of medications are in fact outliers in one’s data, then choosing $\alpha\in\left( 0,1 \right)$ may be more fitting. Note that two people who use a lot of medication may potentially be very far from each other even if their medication use is somewhat comparable as the number of medications itself is penalized. Said in another way, people who use more medication will be further away from everyone when $\alpha\in\left( 0,1 \right)$ as compared with $\alpha\geq1$.

It’s harder to think of scenarios where $\alpha>1$ is appropriate, as it reflects the assumption that people who use more medication ought to be viewed as more central.

The general recommendation is to choose $\alpha=1$ unless the user has strong assumptions about the relevance of medication amounts.

##### Beta

The $\beta\geq1$ parameter allows for the penalization of greater distances. If one is working with a smaller and more homogeneous dataset choosing $\beta>1$ may provide greater opportunity for differentiating between individual medication patterns. However, as dimensions rise with inclusion of more medication types or more different medication types it becomes preferable to choose smaller, if not simply $\beta=1$. A conservative approach would be to keep the parameter equal to 1 in order to avoid the loss of interpretability of proximity under the distance measure (1).

#### Examples of tuning according to context

##### Example 1: Wide selection of ATC codes in a heavy user group

Suppose one is studying a wide range of ATC codes in an older population, who uses a lot of medication. In this example medication from all ATC main groups are studied, and the goal is to model both the therapeutics and indication of the drugs, but also include some modelling of the chemical substance. For this reason, theta values between 1D and 1E in supplementary figure 1 above will be explored.

Moreover, suppose the study period is very long, e.g. from age 60 to 70 with monthly data on percentage of defined daily dose. To keep it simple $p=1$and $\beta=1$ is chosen. In this example, the researchers are not too concerned with the precise timing of the exposure, so they might explore values of $\gamma$ around the value calculated according to formula (1) above. As most people in this group use multiple medications it is undesirable to penalize multiple medication use and therefore $\alpha=1$ is chosen.

As most users in this group use multiple medication from various different medication groups concomitantly it is important to choose an appropriate summary method. If the researchers have the assumptions that there are distinct and important medications patterns involving multiple medications that must be well described, then one should choose the “average” summary method. However, if one wishes to favor the common link between medication profiles the “similar representatives” summary method is favored.

##### Example 2: Narrow selection of ATC codes with emphasis on dose

Suppose one is studying statins (ATC codes C10AA) and wish to model the lipid lowering effects of different dose trajectories. This is a very narrow group of medications, with only 8 distinct ATC codes that only differ on the fifth ATC level. Thus, there are only two modelling strategies: either ATC differences are not modelled at all, $\theta=\left( 0,0,0,0,0,0 \right)$, or they are modelled, $\theta=\left( 1,1,1,1,1,1 \right)$.

If one chooses not to model ATC differences, $\theta=\left( 0,0,0,0,0,0 \right)$, then a classical longitudinal clustering is conducted. In this setup, the Euclidean distance, $p=2$, is traditional. Choosing the “average” summary method and standard values of $\alpha=1$, $\beta=1$ and $\gamma=1$ is preferred to maintain the classical approach.

If one wishes to model ATC differences, $\theta=\left( 1,1,1,1,1,1 \right)$, the choice of $\gamma$ becomes central. Does one want to model ATC code specific trajectories then choosing $\gamma\leq\frac{1}{2}\max_{i,m,t} d_{imt}$ and the “similar representatives” summary method with a larger number of clusters is appropriate. If one wishes to collapse some ATC codes by shared dose trajectory then choosing $\gamma=\max_{i,m,t} d_{imt}$ with the “average” summary method and 5-10 clusters may be a good choice.

Another important aspect of modelling doses, is the dose scale. If we wish to have comparability between ATC codes, then the doses must be on a comparable scale. Fluvastatin (C10AA04) is typically given at a higher dose than Simvastatin (C10AA01), so if they are to be comparable then it may be more useful to model their percentage of Defined Daily Dose, percentage of maximum recommended dose, or percentage of recommended initial dose, than milligrams.

##### The example from the use section in this paper

In the example from this paper we consider a fairly narrow selection of ATC codes, namely the Anti-depressant group N06A consisting of 72 distinct ATC codes, 26 of which are in common use in Denmark. Thus, only the careful tuning of $\theta_{4}$ and $\theta_{5}$ is important, as the other values of theta never become relevant. The values $\theta_{4}=1$ and $\theta_{5}=0.4$ were chosen, which gives a metric that believes that the mode of action is slightly more important than the precise chemical compound of the active ingredient. This choice was made to have a clearer distinction between the SSRI, SNRI and the other anti-depressant groups.

This example examines the pregnancy, a time period where the timing of exposure may be for critical importance, so $\gamma=4$ was chosen. This is four time the balanced gamma proposed in formula (1) above, as it is generally theorized that the precise timing of exposure may be central to the understanding of medication exposure effects.

Standard values of $p=1$, $\alpha=1$ and $\beta=1$ were chosen, as we did not have any strong assumptions on the need for penalizing concomitant medication use or outlier medication profiles.

The studied medication group is quite small and the population typically only uses one of the studied medications. Moreover, we do not believe that complex concomitant medication use should be particularly centered in this study. Therefore, the “similar representatives” summary method was chosen.

Lastly, the Ward linkage (specifically the “ward.D2” linkage) was chosen after comparing and contrasting the resulting constellations learned using both the Ward and complete linkage. The ward linkage was chosen as it produced a constellation with a more uniform distribution of exposed pregnancies into clusters. This more even distribution into clusters was seen as desirable to ensure sufficient power in the following analyses.

[1] Aggarwal CC, Hinneburg A, Keim DA. On the Surprising Behavior of Distance Metrics in High Dimensional Space. In Database Theory — ICDT 2001; 2001; London, UK. p. 420–434.

##### Overview of our suggested tuning strategies

In supplementary table 2 below we have collected the tuning suggestions from the above sections of Supplementary 2 into a reference table.

| **Supplementary Table 2.** An overview of the different challenges and user context and the authors recommended tunings in these scenarios. | | |
| --- | --- | --- |
| **User context, challenges and scenarios** | **General tuning recommendation*** | |
| *Design and clinical interest* |  | |
| Broad categorizations with lower tolerance for spurious finds | Complete linkage & fewer clusters | |
| Balanced approach | Average or Ward linkage | |
| Identification of smaller groups   with moderate allowance for spurious finds | Average or Ward linkage  & more clusters | |
| Identification of smaller groups   with hExplorative study, higher allowance for spurious finds | Average, Ward or single linkage  & more clusters | |
| *The range of the studied medication* | *Distance measure* | |
| A wide range with less comparability | Similar representatives | |
| A narrow range with more comparability | Average | |
| *Relative ATC level importance* | $\theta$ *from Supp. Fig. 1* | |
| Chemical levels of high relative importance | 1A, 1B, 1G | |
| Equal importance of all ATC levels | 1D | |
| Pharmacological or therapeutic levels of high relative importance | 1C, 1H, 1I, 1J, 1K | |
| Pharmacological or therapeutic levels of low relative importance | 1E | |
| Anatomical levels of high relative importance | 1F & 1L | |
| *Balancing ATC and timing importance* | *See formula (1)* | |
| Great timing importance | $\gamma>{\theta_{0}}/{\max_{i,m,t} d_{imt}}$ | |
| Equal importance | $\gamma={\theta_{0}}/{\max_{i,m,t} d_{imt}}$ | |
| Almost no timing importance | $\gamma<{\theta_{0}}/{\max_{i,m,t} d_{imt}}$ | |
| *Importance of dosage or precise timing* |  | |
| A range of doses may be comparable | $p=1$ | |
| High importance of the precise timing or dosage | $p\in(1,2]$ | |
| Very high importance of the exact timing or dosage | $p>2$ | |
| *Does multiple drug use set you apart?* |  | |
| People who use many drugs are like others | $\alpha=1$ | |
| People who use many drugs are different from everybody else | $\alpha>1$ | |
| *Overall heterogeneity in data* |  | |
| There is sufficient heterogeneity in data (this is the most typical case) | | $\beta=1$ |
| Peoples medication use is very similar and all differences must be exaggerated | | $\beta>1$ |
| * These are general recommendations, and researchers should exercise discretion based on the specific characteristics of their dataset and the objectives of their study. The appropriateness of the suggested strategies may vary depending on the nature of the data and the clinical context. | | |

### Supplementary 4. Algorithm structure and time complexity

The algorithm presented in this paper involves three major steps that can cause both the runtime and space requirements to grow as the number of observations studied increase. The first step is to determine the unique medication patterns in the data, the second step constructs the distance matrix, and the final step is the application of the agglomerative hierarchical clustering analysis (HCA).

There is no need to apply the clustering algorithm to people with identical medication profiles, as these people have distance 0 to each other and identical distances to everyone else. Therefore, the first step of the algorithm is to identify unique medication patterns, so we can work with this smaller dataset in the following more time complex steps. This also means that the time complexity of the following steps is guided by the number of unique medication pattern in the data and not the number of observations. Thus, when the data is fairly narrow or homogeneous, this may shave a substantial amount for time from the following steps.

If more than one set of tuning parameters is being explored in one run, then the following steps may be parallelized. In the tame package, parallelization is implemented via the parallel package. As the above step of finding unique medication patterns is always the same for a specific dataset, the user may save some time be setting the parallel in the medic() function to TRUE or the number of desired CPU’s to utilize.

With the reduced set of unique observations, the distance matrix is created. It’s only necessary to compute the lower triangular matrix, as the distance matrix is symmetric. This step has been implemented in C++ to speed up the process significantly.

The time complexity of the standard agglomerative HCA easily derived and is $\mathcal{O(}n^{3})$ where $n$ is the number of unique medication profiles in the data. This is the limiting factor of this entire algorithm. Some agglomerative HCA methods, such as the single linkage and complete linkage methods, can achieve a reduced runtime of $\mathcal{O(}n^{2})$. In this R-package, we use the hclust() function from the stats R-package, which has a very efficient implementation in Fortran.

Below is a simple benchmarking study of subsets of the pregnancy exposure data used as an example in this paper. A number of pregnancies were sampled without replacement from the base cohort, and the medic() algorithm was applied and timed. This was done on a large server with an Intel(R) Xeon(R) Platinum 8176 CPU @ 2.10GHz.

We see that on this system, analyzing anti-depressant use during pregnancy in a cohort of 15,000 pregnancies takes less than 2 minutes, but if the study cohort increases to 30,000 pregnancies, an analysis takes between 5 and 6 minutes. Thus, the computational time increases super-linearly with the number of observations as expected.

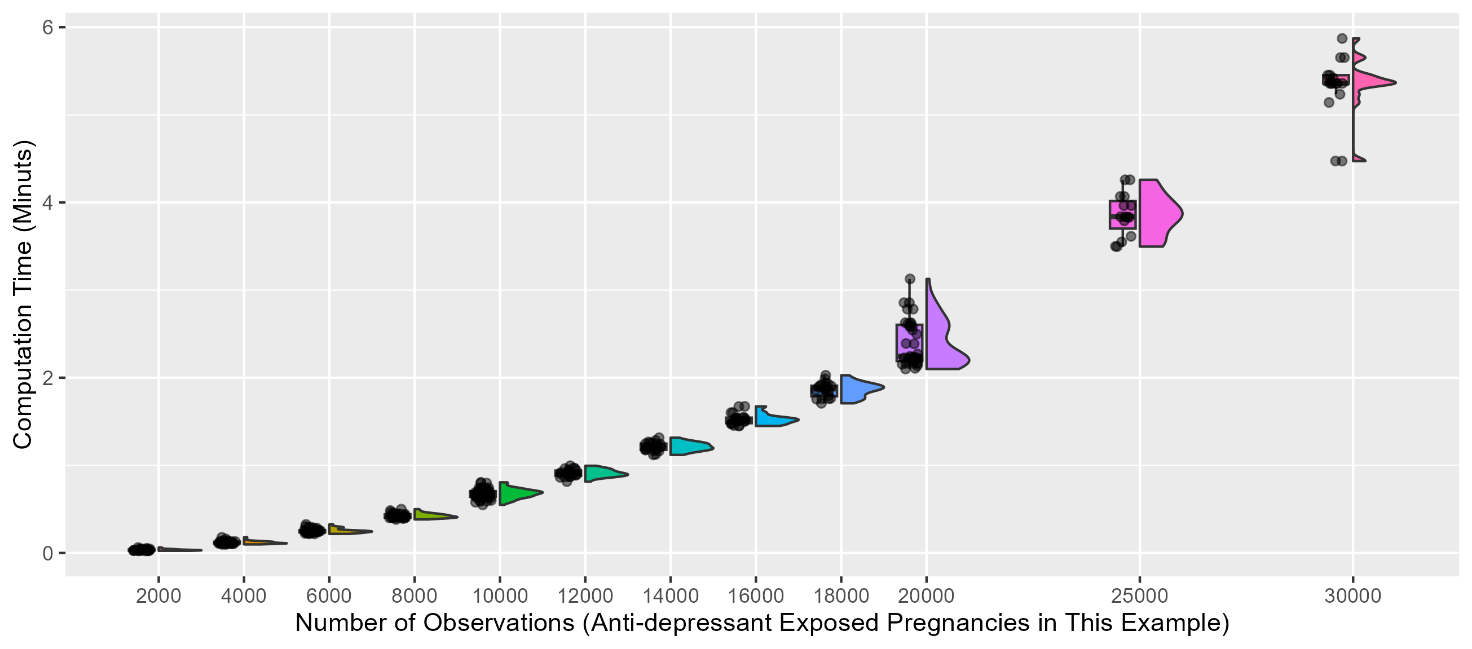

**Supplementary figure 2.** Computational time of medic() clustering of subsets of the pregnancy exposure data from the article use example.

### Supplementary 5. Examples of how to encode data

Here we have created some synthetic data examples to illustrate different ways of encoding data for analysis with medic() in tame.

Supplementary table 2 below illustrates the data format used for the analysis of anti-depressant exposure patterns before and during pregnancy in this paper. In this example medication exposure is binary (exposed / not exposed).

| **Supplementary table 2.** Synthetic data example formatted in the same way as the data analyzed in this paper: exposure yes/no by week from 24 weeks before pregnancy until week 36 of the pregnancy. | | | | | | | | | |
| --- | --- | --- | --- | --- | --- | --- | --- | --- | --- |
| **Id** | **ATC** | **Week -24** | **Week -23** | **…** | **Week 1** | **Week 2** | **Week 3** | **…** | **Week 36** |
| 1 | N06AB04 | 1 | 1 | … | 1 | 1 | 0 | … | 0 |
| 1 | N06AX16 | 0 | 0 | … | 0 | 1 | 1 | … | 1 |
| 2 | N06AB04 | 0 | 0 | … | 1 | 1 | 1 | … | 0 |
| 3 | N06AB03 | 0 | 0 | … | 0 | 0 | 1 | … | 1 |
| 3 | N06AX11 | 1 | 1 | … | 1 | 0 | 0 | … | 0 |
| 3 | N06AA02 | 1 | 1 | … | 1 | 1 | 0 | … | 0 |
| 4 | N06AA10 | 0 | 1 | … | 1 | 1 | 1 | … | 1 |
| 5 | N06AB04 | 1 | 1 | … | 1 | 0 | 0 | … | 0 |
| 5 | N06AB10 | 1 | 1 | … | 1 | 0 | 0 | … | 0 |
| 6 | N06AX16 | 1 | 1 | … | 1 | 1 | 1 | … | 1 |
| $\vdots$ | $\vdots$ | $\vdots$ | $\vdots$ |  | $\vdots$ | $\vdots$ | $\vdots$ |  | $\vdots$ |

The *dose trajectory distance measure* (see supplementary 1) is given as the Minkowski distance, and it is therefore important that the interpretation of dose is consistent within and across the medication types studied. Thus, some type of standardization is typically required. Below, supplementary table 3 and 4 illustrate data with both exposure timing and dose information that solve this dose standardization in two different ways.

In supplementary table 3 WHO’s defined daily dose (DDD) is used to standardize across medication types. As DDD is defined as the assumed average maintenance dose per day for a drug used for its main indication in adults, this method requires that main indications of the medications have comparable levels of medication ‘potency’.

| **Supplementary table 3.** Synthetic data example with both exposure timing and dose information where dose is measured as a percentage of the defined daily dose. | | | | | | | | | |
| --- | --- | --- | --- | --- | --- | --- | --- | --- | --- |
|  |  | **Percentage of defined daily doses ingested per day** | | | | | | | |
| **Id** | **ATC** | **Week 1** | **Week 2** | **Week 3** | **Week 4** | **Week 5** | **Week 6** | **Week 7** | **Week 8** |
| 1 | N06AB04 | 1.0 | 1.0 | 1.0 | 0.8 | 0.6 | 0.4 | 0 | 0 |
| 1 | N06AX03 | 0 | 0 | 0 | 0 | 0.5 | 1.0 | 1.0 | 1.0 |
| 2 | N06AA09 | 1.0 | 1.0 | 1.0 | 1.0 | 1.0 | 1.0 | 1.0 | 1.0 |
| 3 | N06AB04 | 0.8 | 1.0 | 2.0 | 2.0 | 1.8 | 2.0 | 2.0 | 2.0 |
| 4 | N06AB03 | 2.0 | 2.2 | 2.0 | 2.0 | 1.5 | 1.5 | 1.0 | 1.0 |
| 4 | N06AX11 | 0 | 0 | 0.3 | 0.3 | 0.5 | 0.7 | 1.0 | 1.0 |
| $\vdots$ | $\vdots$ | $\vdots$ | $\vdots$ | $\vdots$ | $\vdots$ | $\vdots$ | $\vdots$ | $\vdots$ | $\vdots$ |

In supplementary table 4 the number of pills is used. This method requires that a pill – both within and between medications – is of comparable ‘potency’. Note that it is advisable to choose a small $\gamma$ in the tuning of the distance measure (see supplementary 3), as the size of the doses is large in this example.

| **Supplementary table 4.** Synthetic data example with both exposure timing and dose information where dose is measured as the number of pills redeemed. | | | | | | | | | |
| --- | --- | --- | --- | --- | --- | --- | --- | --- | --- |
|  |  | **Number of pills redeemed within the year** | | | | | | | |
| **Id** | **ATC** | **Year 1** | **Year 2** | **Year 3** | **Year 4** | **Year 5** | **Year 6** | **Year 7** | **Year 8** |
| 1 | N06AB04 | 100 | 300 | 350 | 400 | 200 | 200 | 0 | 0 |
| 1 | N06AX03 | 0 | 0 | 0 | 0 | 150 | 350 | 350 | 350 |
| 2 | N06AA09 | 150 | 150 | 150 | 150 | 150 | 150 | 150 | 150 |
| 3 | N06AB04 | 100 | 150 | 300 | 300 | 250 | 200 | 200 | 150 |
| 4 | N06AB03 | 500 | 600 | 500 | 500 | 300 | 200 | 100 | 100 |
| 4 | N06AX11 | 0 | 0 | 100 | 150 | 300 | 300 | 300 | 300 |
| $\vdots$ | $\vdots$ | $\vdots$ | $\vdots$ | $\vdots$ | $\vdots$ | $\vdots$ | $\vdots$ | $\vdots$ | $\vdots$ |

When only ATC codes is available, such as in supplementary table 5A, the medic() method can still be applied. In this case only the *ATC distance measure* contributes to the distance calculation. If ATC codes and dose is available, but there is no exposure information over time as in supplementary table 5B, one can apply medic() with dose information as a single exposure timepoint.

| **Supplementary table 5A.** Synthetic data example with only ATC codes. | |  | **Supplementary table 5B.** Synthetic data example with dose, but no exposure timing information. | | |
| --- | --- | --- | --- | --- | --- |
| **Id** | **ATC** |  | **Id** | **ATC** | **Average percentage of DDD per day** |
| 1 | N06AA09 |  | 1 | N06AA09 | 95 |
| 2 | N06AB04 |  | 2 | N06AB04 | 60 |
| 2 | N06AX03 |  | 2 | N06AX03 | 30 |
| 3 | N06AB04 |  | 3 | N06AB04 | 80 |
| 3 | N06AB05 |  | 3 | N06AB05 | 10 |
| 3 | N06AX16 |  | 3 | N06AX16 | 25 |
| $\vdots$ | $\vdots$ |  | $\vdots$ | $\vdots$ | $\vdots$ |

Lastly, we consider an example that illustrate the opportunity to study other aspects of medication. In this example self-reported adherence is encoded numerically and may be studies as an “adherence trajectory” using medic().

| **Supplementary table 6.** Synthetic data example with that studies adherence patterns by medication rather than exposure. | | | | | | |
| --- | --- | --- | --- | --- | --- | --- |
|  |  | **Self-reported adherence by medication and month*** | | | | |
| **Id** | **ATC** | **First month** | **Second month** | **Third month** | **Fourth month** | **Fifth month** |
| 1 | N06AB04 | 1 | 0 | 1 | 2 | 2 |
| 1 | N06AX03 | 0 | 0 | 0 | 1 | 1 |
| 2 | N06AA09 | 2 | 3 | 3 | 2 | 2 |
| 3 | N06AB03 | 2 | 2 | 3 | 3 | 2 |
| 3 | N06AX11 | 2 | 2 | 2 | 3 | 3 |
| $\vdots$ | $\vdots$ | $\vdots$ | $\vdots$ | $\vdots$ | $\vdots$ | $\vdots$ |
| * 0 = did not take the medication, 1 = took some of the medication, 2 = took most of the medication,   3 = took all the medication | | | | | | |

### Supplementary 6. Additional analysis information

To conduct an analysis of risk of redeeming psycholeptics within one year of birth given learned medication clusters we identified a number of potential confounders.

Maternal age at conception (<25, 25-29, 30-34, ≥35), maternal parity (0, 1, 2, ≥3), year at pregnancy start (1997-2002, 2003-2007, 2008-2012, 2013-2016), maternal pre-pregnancy BMI (<18.5, 18.5-25, 25-30, 30-35, >35), and maternal smoking status (non-smoker, stopped smoking or smoker) were obtained from the Danish Medical Birth Registry.

Using the Danish National Patient Registry we calculated Charlson comorbidity score (0 or ≥1), and obtained information on history of psychiatric hospitalization (ICD-10 F00-F69 and F80-F99), and history of self-harm (ICD-10 X60-X84, T39, T42, T43, T58, or a hospitalization with primary diagnosis ICD-10 F and secondary diagnosis ICD-10 S51, S55, S59, S61, S65, S69).

From Statistics Denmark, we obtained information on maternal family structure (married, single, or living with partner), maternal employment status (employed, employed in a management position, self-employed, or unemployed and receiving public assistance), maternal level of education (primary, secondary, postsecondary, or vocational school), location of residence in Denmark (capital region, central region, northern region, Zealand, or southern region), disposable household income (quartile 1, 2, 3, or 4), and maternal country of origin (Denmark, Europe (without Denmark), or other).

Redemption of opioid analgesics (ATC code N02A), antiseizure medications (ATC code N03A), antipsychotics (ATC code N05A), benzodiazepine/z-hypnotics (ATC codes N05BA, N05CD, N05CF), or anxiolytics (ATC code N05B except for N05BA) in the 6 months leading up to pregnancy were obtained from the Danish National Prescription Registry.

In supplementary table 3 below the cohort characteristics by variables and medication cluster can be found.

| **Supplementary table 7.** Characteristics of the pregnancies in the study cohort by assigned cluster. | | | | | | | |
| --- | --- | --- | --- | --- | --- | --- | --- |
|  | **I** | **II** | **III** | **IV** | **V** | **VI** | **VII** |
| **Age** |  |  |  |  |  |  |  |
| <25 | 2573 (23%) | 1247 (15%) | 957 (22%) | 694 (19%) | 678 (21%) | 187 (14%) | 153 (12%) |
| 25-29 | 3662 (32%) | 2597 (30%) | 1383 (31%) | 1131 (32%) | 1041 (32%) | 428 (32%) | 352 (28%) |
| 30-34 | 3367 (30%) | 3001 (35%) | 1248 (28%) | 1118 (31%) | 964 (30%) | 431 (33%) | 436 (35%) |
| ≥35 | 1670 (15%) | 1741 (20%) | 830 (19%) | 620 (17%) | 558 (17%) | 278 (21%) | 310 (25%) |
| **Parity** |  |  |  |  |  |  |  |
| 1 | 4992 (44%) | 3791 (44%) | 1864 (42%) | 1646 (46%) | 1457 (45%) | 521 (39%) | 560 (45%) |
| 2 | 3655 (32%) | 3042 (35%) | 1372 (31%) | 1170 (33%) | 928 (29%) | 394 (30%) | 440 (35%) |
| 3+ | 2484 (22%) | 1672 (19%) | 1133 (26%) | 703 (20%) | 824 (25%) | 392 (30%) | 236 (19%) |
| Unknown | 141 (1%) | 81 (1%) | 49 (1%) | 44 (1%) | 32 (1%) | 17 (1%) | 15 (1%) |
| **Birth year** |  |  |  |  |  |  |  |
| 1997-2002 | 2197 (19%) | 519 (6%) | 351 (8%) | 283 (8%) | 380 (12%) | 319 (24%) | 41 (3%) |
| 2003-2007 | 3256 (29%) | 1637 (19%) | 1070 (24%) | 1253 (35%) | 804 (25%) | 299 (23%) | 125 (10%) |
| 2008-2012 | 3702 (33%) | 3622 (42%) | 1861 (42%) | 1507 (42%) | 1120 (35%) | 377 (28%) | 525 (42%) |
| 2013-2016 | 2117 (19%) | 2808 (33%) | 1136 (26%) | 520 (15%) | 937 (29%) | 329 (25%) | 560 (45%) |
| **Household income** |  |  |  |  |  |  |  |
| [0-0.25] | 4813 (43%) | 3272 (38%) | 2009 (45%) | 1466 (41%) | 1467 (45%) | 471 (36%) | 531 (42%) |
| (0.25-0.5] | 3545 (31%) | 2839 (33%) | 1462 (33%) | 1130 (32%) | 1028 (32%) | 478 (36%) | 421 (34%) |
| (0.5-0.75] | 2002 (18%) | 1653 (19%) | 661 (15%) | 656 (18%) | 526 (16%) | 255 (19%) | 202 (16%) |
| (0.75-1] | 912 (8%) | 822 (10%) | 286 (6%) | 311 (9%) | 220 (7%) | 120 (9%) | 97 (8%) |
| **Country of origin** |  |  |  |  |  |  |  |
| Denmark | 10016 (89%) | 7861 (92%) | 3771 (85%) | 3204 (90%) | 2767 (85%) | 1083 (82%) | 1162 (93%) |
| Europe | 589 (5%) | 392 (5%) | 283 (6%) | 181 (5%) | 211 (7%) | 86 (6%) | 47 (4%) |
| Other | 667 (6%) | 333 (4%) | 364 (8%) | 178 (5%) | 263 (8%) | 155 (12%) | 42 (3%) |
| **Family structure** |  |  |  |  |  |  |  |
| Married | 3309 (29%) | 2716 (32%) | 1340 (30%) | 1091 (31%) | 924 (29%) | 488 (37%) | 364 (29%) |
| Single | 3832 (34%) | 2435 (28%) | 1590 (36%) | 1175 (33%) | 1185 (37%) | 362 (27%) | 381 (30%) |
| Cohabitation \w partner | 4131 (37%) | 3435 (40%) | 1488 (34%) | 1297 (36%) | 1132 (35%) | 474 (36%) | 506 (40%) |
| **Education** |  |  |  |  |  |  |  |
| Primary | 4088 (36%) | 2298 (27%) | 1750 (40%) | 1150 (32%) | 1236 (38%) | 428 (32%) | 334 (27%) |
| Secondary | 1409 (12%) | 1124 (13%) | 562 (13%) | 530 (15%) | 391 (12%) | 157 (12%) | 171 (14%) |
| Vocational | 2972 (26%) | 2206 (26%) | 1178 (27%) | 889 (25%) | 863 (27%) | 404 (31%) | 348 (28%) |
| Postsecondary | 2702 (24%) | 2896 (34%) | 886 (20%) | 968 (27%) | 706 (22%) | 323 (24%) | 388 (31%) |
| Unknown | 101 (1%) | 62 (1%) | 42 (1%) | 26 (1%) | 45 (1%) | 12 (1%) | 10 (1%) |
| **Emplyment Status** |  |  |  |  |  |  |  |
| Employed | 5260 (47%) | 3908 (46%) | 1715 (39%) | 1623 (46%) | 1420 (44%) | 550 (42%) | 497 (40%) |
| Employed, management | 842 (7%) | 973 (11%) | 262 (6%) | 312 (9%) | 207 (6%) | 104 (8%) | 118 (9%) |
| Self-employed | 164 (1%) | 127 (1%) | 70 (2%) | 58 (2%) | 40 (1%) | 30 (2%) | 19 (2%) |
| Unemployed, public assistance | 5006 (44%) | 3578 (42%) | 2371 (54%) | 1570 (44%) | 1574 (49%) | 640 (48%) | 617 (49%) |
| **Residency region** |  |  |  |  |  |  |  |
| Capital Region | 3164 (28%) | 2486 (29%) | 1012 (23%) | 968 (27%) | 734 (23%) | 318 (24%) | 294 (24%) |
| Central Region | 2792 (25%) | 2502 (29%) | 1246 (28%) | 1079 (30%) | 825 (25%) | 359 (27%) | 319 (25%) |
| Northern Region | 1117 (10%) | 768 (9%) | 400 (9%) | 249 (7%) | 342 (11%) | 133 (10%) | 174 (14%) |
| Southern Region | 2608 (23%) | 1891 (22%) | 1153 (26%) | 823 (23%) | 828 (26%) | 357 (27%) | 295 (24%) |
| Zealand Region | 1591 (14%) | 939 (11%) | 607 (14%) | 444 (12%) | 512 (16%) | 157 (12%) | 169 (14%) |
| **History of use of various drug classes** | | | | | | | |
| Anti-psychotic | 1240 (11%) | 1607 (19%) | 1079 (24%) | 642 (18%) | 588 (18%) | 187 (14%) | 477 (38%) |
| Anti-seizure | 509 (5%) | 831 (10%) | 547 (12%) | 298 (8%) | 318 (10%) | 186 (14%) | 285 (23%) |
| Anxiolytics | 150 (1%) | 195 (2%) | 113 (3%) | 60 (2%) | 76 (2%) | 23 (2%) | 29 (2%) |
| Benzodiazepine | 3899 (35%) | 3920 (46%) | 2186 (49%) | 1598 (45%) | 1329 (41%) | 486 (37%) | 756 (60%) |
| Opiod | 2483 (22%) | 2299 (27%) | 1549 (35%) | 894 (25%) | 962 (30%) | 608 (46%) | 423 (34%) |
| Psycholeptics | 4492 (40%) | 4541 (53%) | 2516 (57%) | 1830 (51%) | 1558 (48%) | 554 (42%) | 885 (71%) |
| **Charlson comordibity score ≥1** | 574 (5%) | 494 (6%) | 294 (7%) | 190 (5%) | 186 (6%) | 93 (7%) | 77 (6%) |
| **History of psychiatric hospitalizations** | 952 (8%) | 1206 (14%) | 770 (17%) | 470 (13%) | 444 (14%) | 97 (7%) | 371 (30%) |
| **History of self-harm** | 712 (6%) | 637 (7%) | 467 (11%) | 286 (8%) | 321 (10%) | 64 (5%) | 156 (12%) |

### Supplementary 7. Output summarizing methods

A complete documentation of the output summarization method can be found in the R-package documentation [1]. As described in the documentation the following are the methods implemented as of September 2024:

- ’cluster_frequency’: The number of individuals assigned to each cluster and the associated frequency of assignment.
- ’medication_frequency’: The number of individuals with a specific ATC code within a cluster. Moreover, it calculates the percentage of people with this medication assigned to this cluster and the percent of people within the cluster who use this medication.
- ‘comedication_count’: The number of ATC codes an individual has, followed by the number of individuals within a cluster that has that many ATC codes. Additionally, various relevant percentages or calculated. See the Value section in the package documentation for more details on these percentages.
- ‘timing_trajectory’: The number of unique timing trajectories in each cluster, and the average timing trajectories in each cluster.
- ‘timing_atc_group’: The number of people with unique timing trajectory and ATC group, as given by a user defined list of ATC code groupings in each cluster.
- ‘all’: All of the above are performed.”

[1] Laksafoss A. tame: Timing, Anatomical, Therapeutic and Chemical Based Medication Clustering. <https://cran.r-project.org/web/packages/tame/tame.pdf>

### Supplementary 8. Full Cluster Characterization

In the analysis conducted for illustration of the method, we chose to learn seven clusters using the Ward linkage, the similar representatives distance measure, and the following tuning parameters:

$\alpha=1$, $\beta=1$, $\gamma=4$, $\theta=\left( 1, 1, 1, 1, 0.4, 0 \right)$ and $p=1$.

A discussion on how to tune these parameters and why these parameters were chosen can be found in S2.

#### Cluster I – “Discontinuation of SSRIs”

The largest cluster, with 33% of pregnancies, consists almost exclusively of SSRI single drug users, who discontinue their anti-depressant during the pregnancy. Less than 3% of pregnancies are exposed to any anti-depressant by week 20, and more than 99% of these pregnancies are only exposed to 1 anti-depressant. Almost half, 48%, of the pregnancies are exposed to the SSRI N06AB04.

#### Cluster II – “Sustained use of one SSRI”

Similar to cluster I, the second cluster, with 26% of pregnancies, is almost exclusively SSRI single drug users. However, the pregnancies in this cluster has a high level of sustained SSRI usage with at least 70% of pregnancies being exposed at any time point in the study period, and with more than 90% of pregnancies being exposed in the weeks 0 to 13.

#### Cluster III – “Concomitant use of different types of antidepressants”

The third largest cluster with 13% of pregnancies studied, is characterized by concomitant drug use of a mix of all 3 classes of anti-depressants. More than 88% of pregnancies are exposed to either SSRI or SNRI or both, while only 18% of pregnancies are exposed to other anti-depressants. The SSRI use frequency remains fairly stable across the entire exposure period, as opposed to the SNRI use frequency which drops at the start of the pregnancy.

#### Cluster IV – “Multiple SSRIs used”

Cluster IV, which consists of 11% of pregnancies is characterized by multiple drug use, with less than 3% using only one drug, almost 88% using 2 anti-depressants, and 10% using 3 or more anti-depressants. Unlike cluster III, where pregnancies are exposed to a mix of different types of antidepressants, in this cluster, the exposure is with multiple SSRIs.

#### Cluster V – “Discontinuation of SNRIs”

Cluster V has 10% of pregnancies and is dominated by single drug SNRI users who discontinue their usage early in the pregnancy. Fewer than 5% of pregnancies are exposed to any anti-depressant by week 20 in this cluster. More than 97% of pregnancies are exposed to only one anti-depressant in this cluster. Various different SNRIs are used in this cluster, but the SNRI N06AX16 is the most commonly used (39%).

#### Cluster VI – “Single use of antidepressants in the other anti-depressants group”

The second smallest cluster, with 4% of pregnancies, is the only cluster dominated by use of other anti-depressants with 100% pregnancies being exposed to an antidepressant in the other anti-depressant group. Use of anti-depressants is more sporadic in this cluster as compared with many of the other clusters. At any given time point in the exposure period, less than 31% of pregnancies are exposed to an anti-depressant, and by week 20 less than 5% of pregnancies are exposed to an anti-depressant. Various antidepressant from the other antidepressant group is used, but the ATC code N06AA09 is the most commonly used (64%).

#### Cluster VII – “Sustained use of SNRI or other-antidepressants”

The smallest cluster, with only 4% of studied pregnancies, is characterized by a sustained use of SNRIs or antidepressant in the other-antidepressant group. A large majority, 75%, of pregnancies are exposed to the SNRI N06AX16. At any given time in the exposure time period at least 80% of pregnancies are exposed to an anti-depressant, with at least 90% of pregnancies being exposed in weeks 0 to 12.
